# Study protocol for a randomized, double-blind, placebo-controlled clinical trial to assess chemoprotective efficacy of Aqueous standardized extract of *Tinospora cordifolia* (AsSET) in patients receiving Docetaxel based chemotherapy for advanced / metastatic solid tumors

**DOI:** 10.64898/2026.10.29.26366795

**Authors:** Priyanka Singh, Aditi Shirsat, Shomita Shetty, Purva Bhor, Amit Joshi, ADB Vaidya, Nirmala Rege, Kumar Prabhash, Vanita Noronha, Anant Ramaswamy, Vikas Otswal, Ashwini Raut, Jayashree Joshi, Vikram Gota

## Abstract

**Introduction:** Docetaxel is widely used for advanced and metastatic solid tumours, but dose limiting toxicities, particularly haematological adverse events, often lead to treatment delays, dose reductions or discontinuation and impair quality of life. Tinospora cordifolia (Guduchi), long used in Ayurveda for its immunomodulatory and rejuvenating properties, has shown chemoprotective potential in experimental and early clinical studies, but robust randomized evidence is lacking. This trial evaluates whether adding an aqueous standardized extract of T. cordifolia(AsSET) to docetaxel-based chemotherapy reduces grade ≥2 adverse events and improves quality of life in patients with advanced or metastatic solid tumours.

**Methods and analysis:** This randomized, double-blind, and placebo-controlled trial designed to evaluate the chemoprotective efficacy of an aqueous standardised extract of Tinospora cordifolia (AsSET) in patients with advanced, recurrent or metastatic solid tumors receiving docetaxel-based chemotherapy. A total of 81 eligible patients that have provided informed consent were recruited in the study. Participants were randomized 2:1 to AsSET or placebo using stratified permuted block randomization stratified by planned cumulative docetaxel dose (≤ 225 vs >225 mg/m^2^) except five participants enrolled before that date were allocated by minimization. All participants received standard intravenous docetaxel-based chemotherapy plus oral administration of either AsSET (two 300mg tablets, 600mg, twice daily) or matching placebo, from enrolment until completion of docetaxel-based therapy. Clinical, laboratory, imaging, and questionnaire based-assessment were performed at baseline, at scheduled interval during treatment and at the final visit. In consenting participants, blood samples for pharmacokinetics were collected once, at nine timepoints up to 24 hours, in any chemotherapy cycle from cycle 2 onwards. Adverse events were graded according to CTCAE version 5.0. The primary outcome is the proportion of patients experiencing grade ≥2 adverse events from first dose to study completion (up to 24 weeks). Secondary outcomes include quality of life measured by the EORTC QLQ-C30 and the disease-site-specific FACT scale, progression-free survival (PFS) per RECIST v1.1, haematological and biochemical parameters, and pharmacokinetic parameters. The database was locked on 17 September 2026 and the locked dataset was transferred to an independent statistician who remains blinded to treatment allocation. Unblinding has not yet been done and outcome analysis has not started.

**Ethics and dissemination:** This study was approved by the Institutional Ethics Committee III (IEC) of the Advanced Centre for Treatment Research & Education in Cancer, Tata Memorial Centre, (ACTREC), Kharghar, Sector 22, Navi Mumbai (project approval no. 900950; first approved on 28 June 2023; this paper describes the most recently amended protocol, version 4.0 dated 24 February 2026; amendments are listed in Annexure 1). The trial is registered with the Clinical Trials Registry-India (CTRI/2023/08/056129) and is conducted in accordance with ICH Good Clinical Practice, the new drugs and Clinical Trials Rules 2019. All participants provided written informed consent, with an additional consent for pharmacokinetic sampling. Participants data are coded to protect confidentiality, and no participant will be identified in any publication without permission. The findings from this study will be presented at an academic conference and published in scholarly journals regardless of the findings.

Trial registration: CTRI/2023/08/056129

**Strengths and limitation of this study:**

□ Robust design in the form of randomized, double-blind, placebo-controlled trial with stratified block randomization provides good internal validity
□ The study is adequately powered to demonstrate the treatment effect for the underlying assumptions
□ Pharmacokinetic sampling in a subset of participants allows assessment of whether AsSET alters docetaxel exposure, an important concern for herb-drug interactions in integrative oncology.
□ The study, being limited to docetaxel-based chemotherapy regimens at a single institution, may not be generalizable to other cytotoxic chemotherapy regimen.
□ The inclusion of patients with multiple solid tumours receiving varied docetaxel-based regimens may lead to heterogeneity in the chemotherapy-related adverse events which could affect the interpretability of pooled results.
□ The 2:1 allocation reduces statistical efficiency compared with equal allocation.

## INTRODUCTION

### Background and rationale

Cancer is a leading cause of mortality worldwide, with close to 20 million new cases and an estimated 9.7 million deaths in 2022. (1). In India, an estimated 1,461,427 new cancer cases occured in 2022, one in nine people is likely to develop cancer in their lifetime, and incidence is expected to rise by 12.8% by 2025 compared with 2020. Males majorly develop lung cancer, while leading cases in females are breast cancer (2). The leading sites overall include cancer of the breast, lung, mouth, cervix uteri, and tongue (3)

Cancer treatment includes surgery, radiotherapy, immunotherapy and systemic chemotherapy, often in combination. Chemotherapy remains a cornerstone of systemic treatment, especially for advanced and metastatic disease, where local treatments alone are insufficient. However, chemotherapy-related adverse events often lead to treatment interruption or discontinuation, which may compromise quality of life (QoL) (4). Common effects include decreased appetite, fatigue, nausea, vomiting and oral mucositis (5). Chemotherapy-induced peripheral neuropathy is also associated with substantial morbidity, including depression, insomnia and ataxia [6].

Docetaxel is a taxane-class drug approved by the US Food and Drug administration for several solid tumours, including metastatic breast, non-small cell lung, prostate, gastric and head and neck cancers (7). Despite its established efficacy, it causes substantial haematological toxicity, with neutropenia and anaemia among the most frequent adverse events (7). Non-haematological toxicities such as fatigue, peripheral neuropathy, mucositis and fluid retention also impair health and QoL, (5,25). These toxicities highlight an unmet need for adjunctive therapies that reduce toxicity without compromising the efficacy of chemotherapy.

Tinospora cordifolia (Guduchi or Giloy), is a widely used herb in Ayurvedic medicine with a long history of application in traditional Indian literature (8). It is known for its immunomodulatory, antioxidant, anti-inflammatory and cytoprotective properties(8,9)(Figure 1). The active chemical constituents of T. cordifolia span several classes, including alkaloids, glycosides, steroids, polysaccharides(10). Previous studies with Guduchi have demonstrated stimulation of granulocyte-macrophage colony-stimulating factor (GM-CSF) enhancing leukocyte counts, and preventing chemotherapy-induced myelosuppression (11) and T. cordifolia induced colony-stimulating activity in serum (12). In humans, a double-blind placebo-controlled study in patients with chronic renal failure reported in an MSc thesis, found higher GM-CSF levels with T. cordifolia (13), and its use in burn management was associated with a significant rise in white cell count and IgG (14).

**Figure 1:**
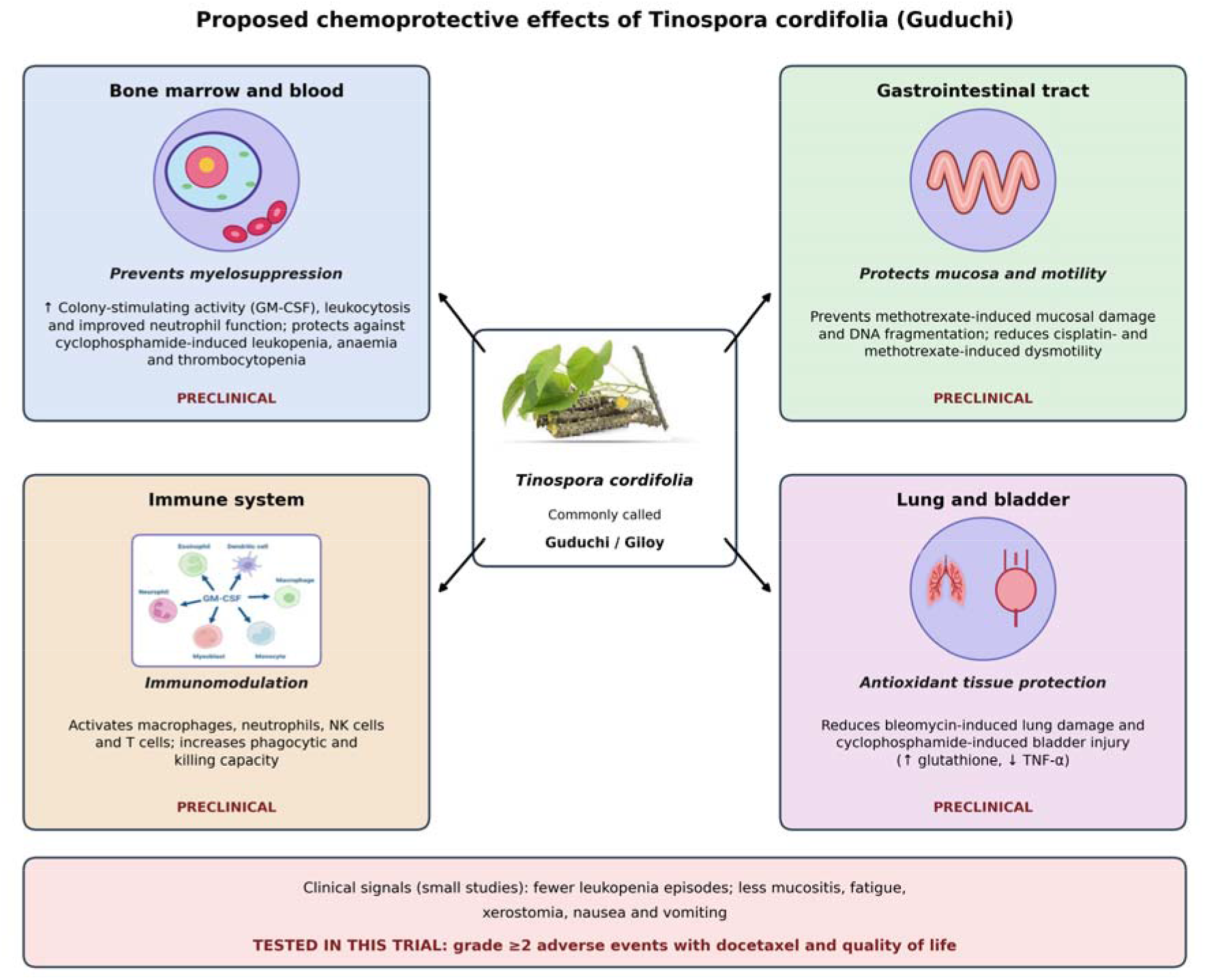
Proposed chemoprotective effects of Tinospora cordifolia.

The most directly relevant clinical evidence comes from two small studies. In a placebo-controlled study of women with breast cancer receiving CMF chemotherapy, an aqueous extract of T. cordifolia (1500 mg/day), started seven days before the first cycle and continued through six cycles, was associated with less leukopenia (WBC <2000/cmm): 11 of 46 analysable cycles with the extract versus 24 of 52 with placebo (p = 0.03). Neutropenia in the first three cycles was also significantly less frequent with the extract, while the more frequent fall of ANC below 500/cmm with placebo was not statistically significant (15).

More recently, Beriwal et al. evaluated Guduchi-ghana (a solidified aqueous extract of T. cordifolia; 1 g tablets three times daily for three months) in patients with cancer who had chemotherapy-related adverse effects (16). Treated patients showed reductions in mucositis, fatigue, xerostomia, nausea-vomiting, skin reactions and tastelessness, with the least effect on alopecia, and haemoglobin, leucocyte and platelet counts improved in 68% of treated patients compared with controls. However, the study was small (56 patients in four groups), did not report actual haematological values, analysed groups before and after treatment without between-group comparison, and did not follow CONSORT reporting guidance.

Taken together, these findings suggest that T. cordifolia may attenuate chemotherapy-related adverse effects, but the evidence comes from small, methodologically limited studies. To our knowledge, no adequately powered, prospectively registered randomized trial has evaluated a standardised T. cordifolia extract as an adjunct to docetaxel-based chemotherapy. This trial was designed to address that gap by testing whether adding an aqueous standardised extract of T. cordifolia (AsSET) to docetaxel-based chemotherapy reduces grade ≥2 adverse events and improves quality of life.

### Objectives

This randomized, double-blind, placebo-controlled trial aims to evaluate whether adding a standardized aqueous extract of Tinospora cordifolia (AsSET) to docetaxel-based chemotherapy reduces chemotherapy-related adverse events in patients with advanced, recurrent or metastatic solid tumours.

The primary objective is to compare the incidence of treatment-emergent grade ≥2 adverse events (haematological and non-haematological) between docetaxel-based chemotherapy with add-on AsSET and docetaxel-based chemotherapy with add-on placebo.

The secondary objectives are to compare the two arms with respect to: (i) general and cancer-specific quality of life (QoL); (ii) progression-free survival (PFS) per RECIST v1.1 (iii) haematological and biochemical parameters and (iv) docetaxel pharmacokinetic parameters. We hypothesised that add-on AsSET would reduce the proportion of participants with grade ≥2 adverse events compared with placebo.

### Trial design

The study is a randomized, double blind, placebo-controlled trial designed to evaluate the safety and efficacy of AsSET. It employs a parallel-group design with an unequal allocation ratio of 2:1. The protocol follows the SPIRIT 2025 reporting guidance(17).

## METHODS

### Study design

The trial was conducted at ACTREC-Tata Memorial Centre, Navi Mumbai and Tata Memorial Hospital, Mumbai, with an interventional arm and a control arm. Participants took the study drug orally twice daily from the day of enrolment until completion of docetaxel-based chemotherapy, so the duration of intervention varied with each participant’s chemotherapy schedule; adverse events were followed for up to 24 weeks. The planned study duration was 30 months (24 months of accrual and 6 months of analysis); enrolment ran from 2 November 2023, after the first approval of the Institutional Ethics Committee, to 21 April 2026. Accrual exceeded the planned 24 months because several other trials were recruiting from the same departments in parallel; the Institutional Ethics Committee was kept informed through regular continuing review reports.

### Recruitment

Patients attending the outpatient departments of the participating institutes who had advanced, recurrent or metastatic solid tumours and were scheduled to start docetaxel, alone or combined with other standard chemotherapeutic drugs, were screened and enrolled. The clinical site coordinators explained the study procedures, risks, benefits and duration, and written informed consent was obtained before enrolment.

### Eligibility criteria

Patients’ participation in the trial will undergo screening assessment according to the following set of inclusion and exclusion criteria for the trial.

Changes to eligibility during the trial: the original protocol restricted enrolment to patients with recurrent or metastatic squamous cell carcinoma of the head and neck, non-small cell lung cancer or metastatic castration-resistant prostate cancer who were chemotherapy-naive for advanced disease, and required COVID-19 vaccination. To speed accrual, and because most of the population had by then been vaccinated, the protocol was amended (Annexure 1) to include any advanced, recurrent or metastatic solid tumour, to allow prior chemotherapy other than docetaxel, and to remove the COVID-19 vaccination and testing requirements. These amendments took effect on 14 November 2023 (protocol version 3.1); five participants had been enrolled before that date.

### Inclusion Criteria

□ Written informed consent given voluntarily.
□ Histologically confirmed solid malignancy (advanced, recurrent or metastatic) with evaluable disease, scheduled to receive docetaxel-based chemotherapy.
□ Age 18–65 years (inclusive), male or female
□ ECOG performance status ≤2.
□ Adequate bone marrow reserve platelet ≥100,000/mm³, WBC ≥3000/mm^3^, neutrophil count ≥2000/mm^3^, haemoglobin ≥8.0 g/dL), renal function (GFR ≥50 mL/min) and liver function (total bilirubin ≤2× upper limit of normal [ULN]; transaminases ≤2.5× ULN, or ≤5× ULN with liver metastases and/or obstructive jaundice).
□ Life expectancy of at least 12 weeks.
□ Willingness to practise birth control during treatment and for 2 months afterwards.
□ Negative tests for HIV, HBsAg and HCV.

### Exclusion Criteria

□ Major illness, including active cardiac, hepatic, endocrine, renal or psychiatric disorders not adequately controlled with current therapy.
□ Brain metastases or primary CNS malignancy.
□ Any condition, including laboratory abnormalities, that would place the participant at unacceptable risk, obscure the evaluation of toxicity or alter drug metabolism.
□ Anti-tumour treatment (radiotherapy, targeted therapy, immunotherapy or other investigational drugs) within 4 weeks before the first dose of study drug.
□ Traditional medicines or natural products with anti-tumour indications and/or potential to relieve chemotherapy toxicity within 4 weeks before the first dose.
□ Corticosteroids (prednisone >10 mg/day or equivalent) or other immunosuppressive therapy within 4 weeks before the first dose.
□ Live attenuated vaccine within 4 weeks before the first dose, or planned during the study.
□ Major surgery within 4 weeks before the first dose, or elective surgery planned during the study.
□ Patients who have a history of prior docetaxel use are excluded from the trial.
□ Pregnancy or lactation.

### Interventions

#### Interventional arm

Participants received standard intravenous docetaxel-based chemotherapy (docetaxel alone or with other standard anticancer drugs, at the dose prescribed by the treating physician) plus oral AsSET tablets supplied by Pharmanza Herbal Pvt. Ltd. against a certificate of analysis. Each tablet contained 300 mg of aqueous standardized T. cordifolia extract. Participants took two tablets (600 mg) twice daily (1200 mg/day), 12-hourly after food with water, including on docetaxel infusion days. This dose lies within the range used in earlier clinical studies of aqueous T. cordifolia extracts (15,16,18).

#### Product details

AsSET tablets were round, orange, film-coated tablets, each containing 300 mg of an aqueous extract of the stem of Tinospora cordifolia (wild-crafted, India; extraction solvent water; herb-to-extract ratio 10:1). According to the manufacturer’s specification, the extract contained not less than 5% total bitters, 20% total polysaccharides and 0.7% of the sum of four marker compounds (magnoflorine, 20-β-hydroxyecdysone, tinosporide and columbin), and the specification included an HPLC identification test. All participants received AsSET from a single batch (PHPL/RD/094/CT-102-01), manufactured on 26 August 2023 by Pharmanza Herbal Pvt. Ltd., Gujarat, India. A herbarium voucher specimen of the raw plant material was not available to the study team.

#### Control arm

Participants received the same chemotherapy plus placebo tablets, also supplied by Pharmanza, identical to AsSET tablets in colour, size, shape, texture, smell and taste. Each placebo tablet contained 230 mg of starch (IP) with excipients, and all placebo came from a single batch (PHPL/RD/094/CT-103-01) manufactured on 26 August 2023. The initial shelf life of the study medication (AsSET and placebo) was 3 years from manufacture. When this ended in August 2026, the manufacturer reported that it had re-tested the batches and extended the shelf life by one year, and the same batches continued to be dispensed.

#### Dose modification and interruption

AsSET and placebo doses were not modified for the duration of the study. The dose of docetaxel, other chemotherapy and supportive drugs was individualized according to the usual schedule for each drug. AsSET or placebo could be temporarily held on the advice of the treating or Ayurvedic physician.

#### Concomitant therapy

Antiemetic prophylaxis and other supportive medications (including for infusion or hypersensitivity reactions) followed institutional guidelines, and medications for associated illnesses were allowed. No list of prohibited concomitant herbal products was specified; all concomitant medications were recorded in the case report form at every visit.

Participant timeline and schedule of procedures

Table 1 shows the schedule of enrolment, interventions and outcome assessments;

**Table 1:**
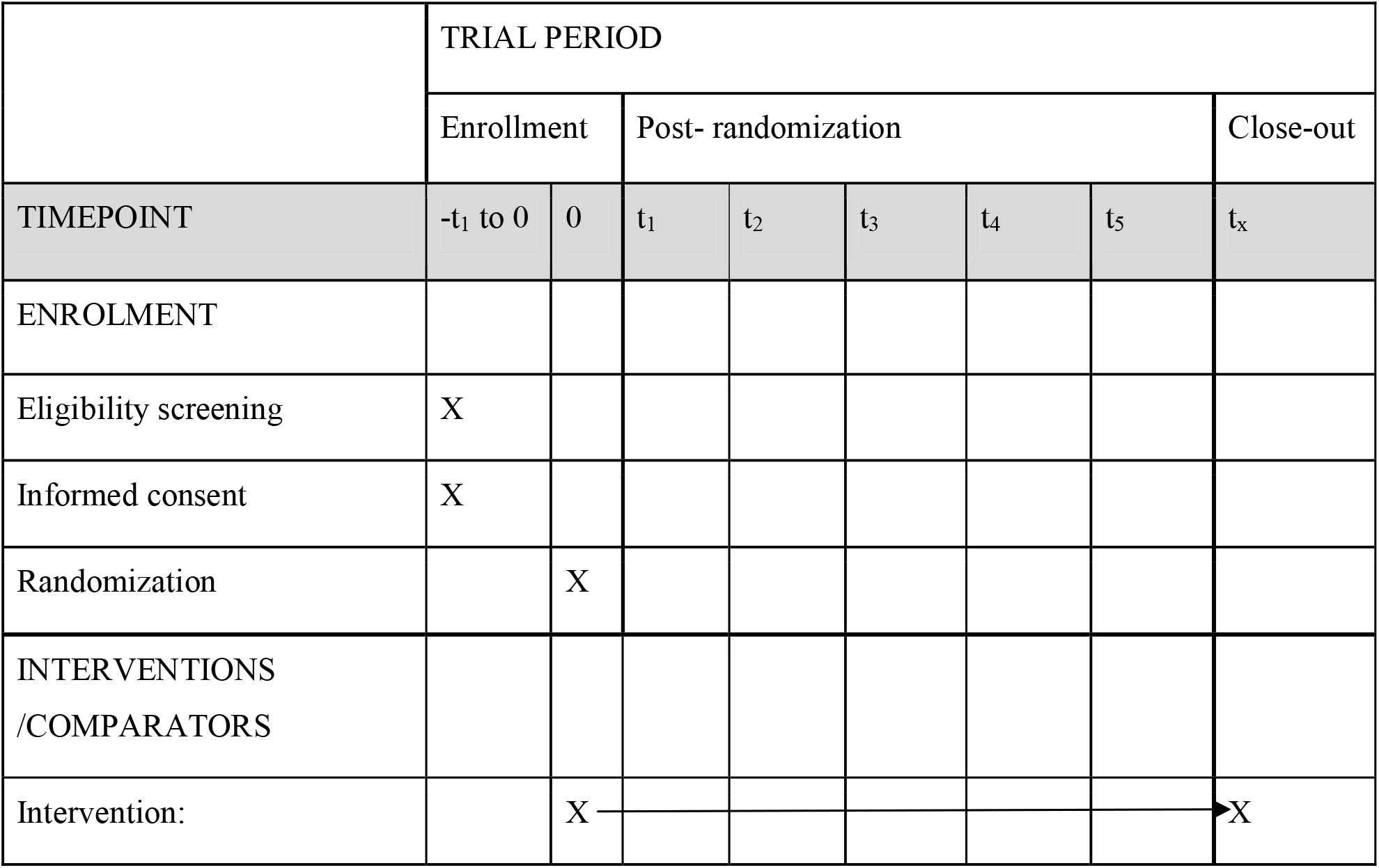
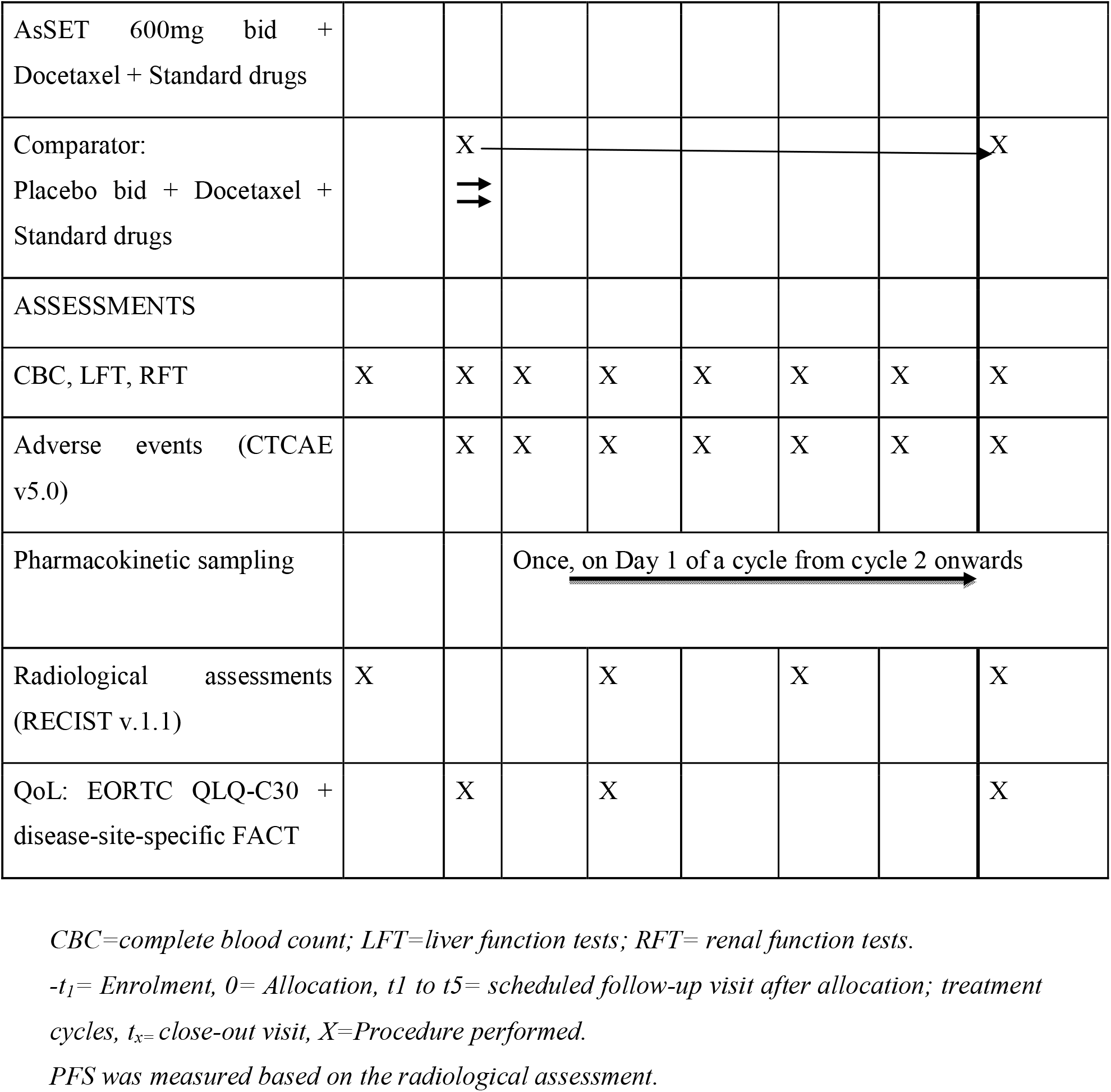
Schedule of enrolment, interventions and outcome assessments.

|  | TRIAL PERIOD |  |  |  |  |  |  |  |
|---|---|---|---|---|---|---|---|---|
|  | Enrollment |  | Post- randomization |  |  |  |  | Close-out |
| TIMEPOINT | -t<sub>1</sub> to 0 | 0 | t<sub>1</sub> | t<sub>2</sub> | t<sub>3</sub> | t<sub>4</sub> | t<sub>5</sub> | t<sub>x</sub> |
| ENROLMENT |  |  |  |  |  |  |  |  |
| Eligibility screening | X |  |  |  |  |  |  |  |
| Informed consent | X |  |  |  |  |  |  |  |
| Randomization |  | X |  |  |  |  |  |  |
| INTERVENTIONS /COMPARATORS |  |  |  |  |  |  |  |  |
| Intervention: |  | X |  |  |  |  |  | X |
| AsSET 600mg bid + Docetaxel + Standard drugs |  |  |  |  |  |  |  |  |
| Comparator:<br>Placebo bid + Docetaxel + Standard drugs |  | X<br>⇉ |  |  |  |  |  | X |
| ASSESSMENTS |  |  |  |  |  |  |  |  |
| CBC, LFT, RFT | X | X | X | X | X | X | X | X |
| Adverse events (CTCAE v5.0) |  | X | X | X | X | X | X | X |
| Pharmacokinetic sampling |  |  | Once, on Day 1 of a cycle from cycle 2 onwards |  |  |  |  |  |
| Radiological assessments (RECIST v.1.1) | X |  |  | X |  | X |  | X |
| QoL: EORTC QLQ-C30 + disease-site-specific FACT |  | X |  | X |  |  |  | X |

*CBC=complete blood count; LFT=liver function tests; RFT= renal function tests.*
*-t<sub>1</sub>= Enrolment, 0= Allocation, t1 to t5= scheduled follow-up visit after allocation; treatment cycles, t<sub>x=</sub> close-out visit, X=Procedure performed.*
*PFS was measured based on the radiological assessment.*

Table 2 shows the study procedures at each visit.

**Table 2:**
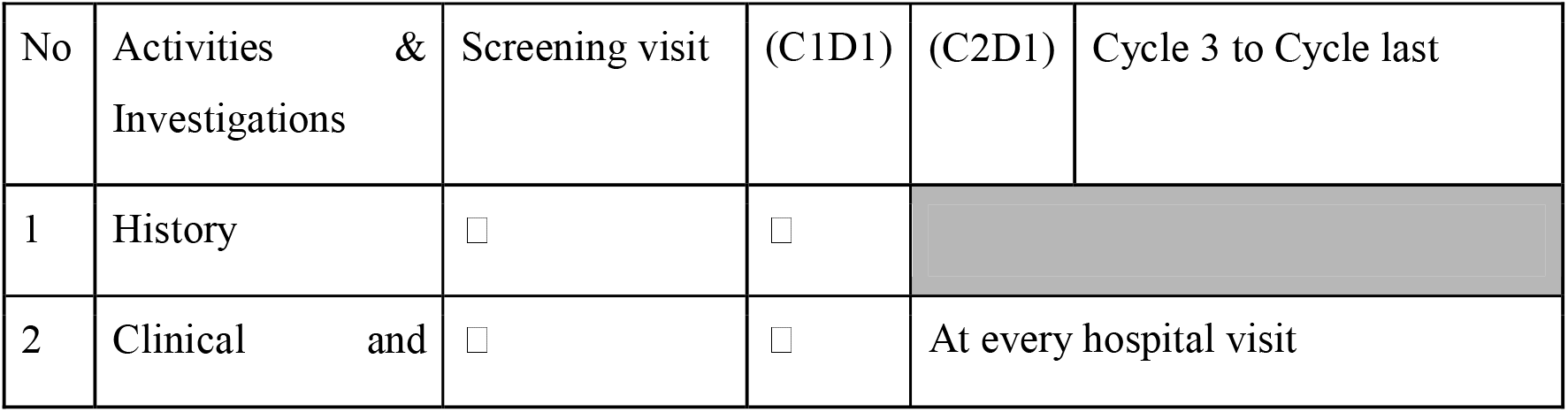
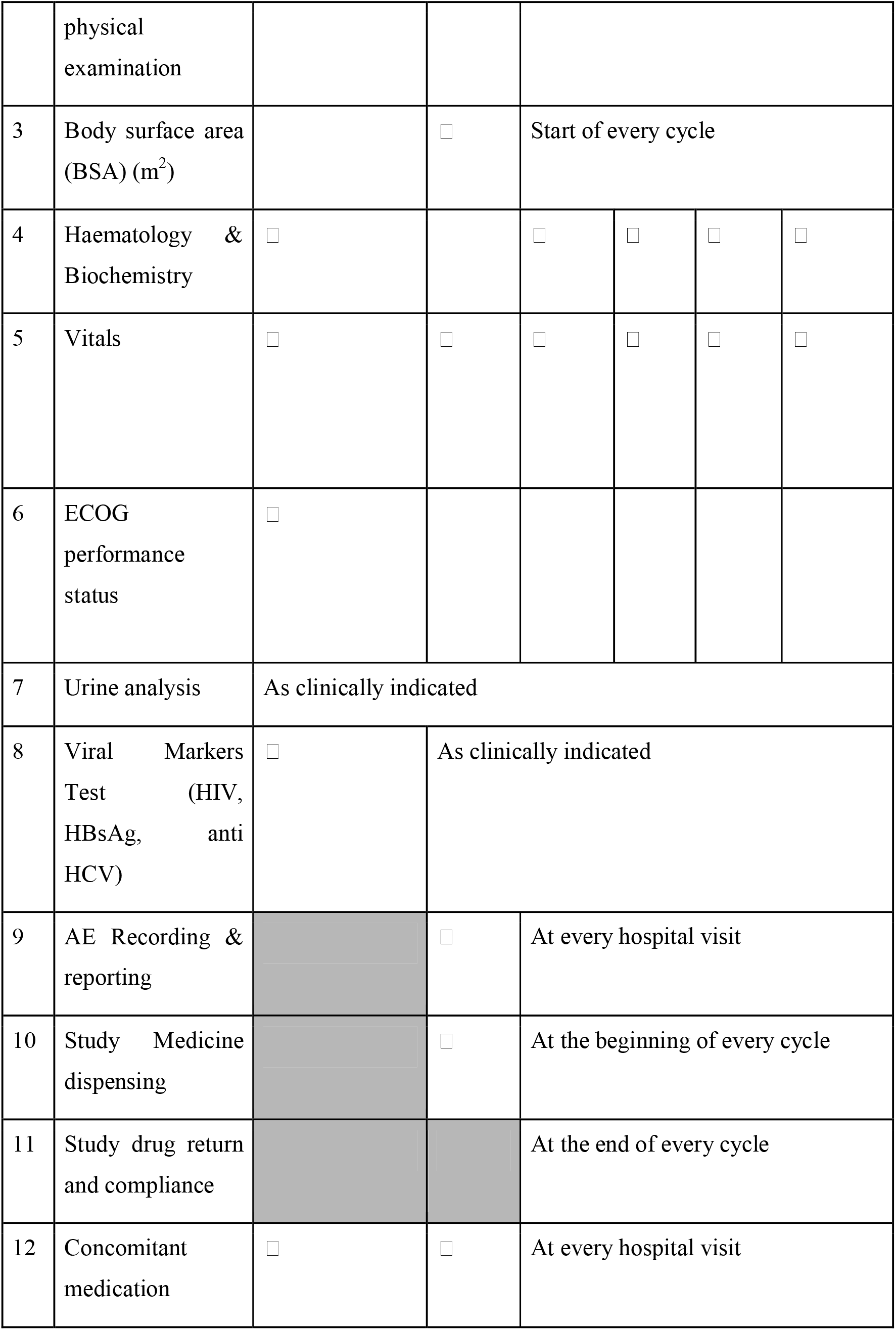
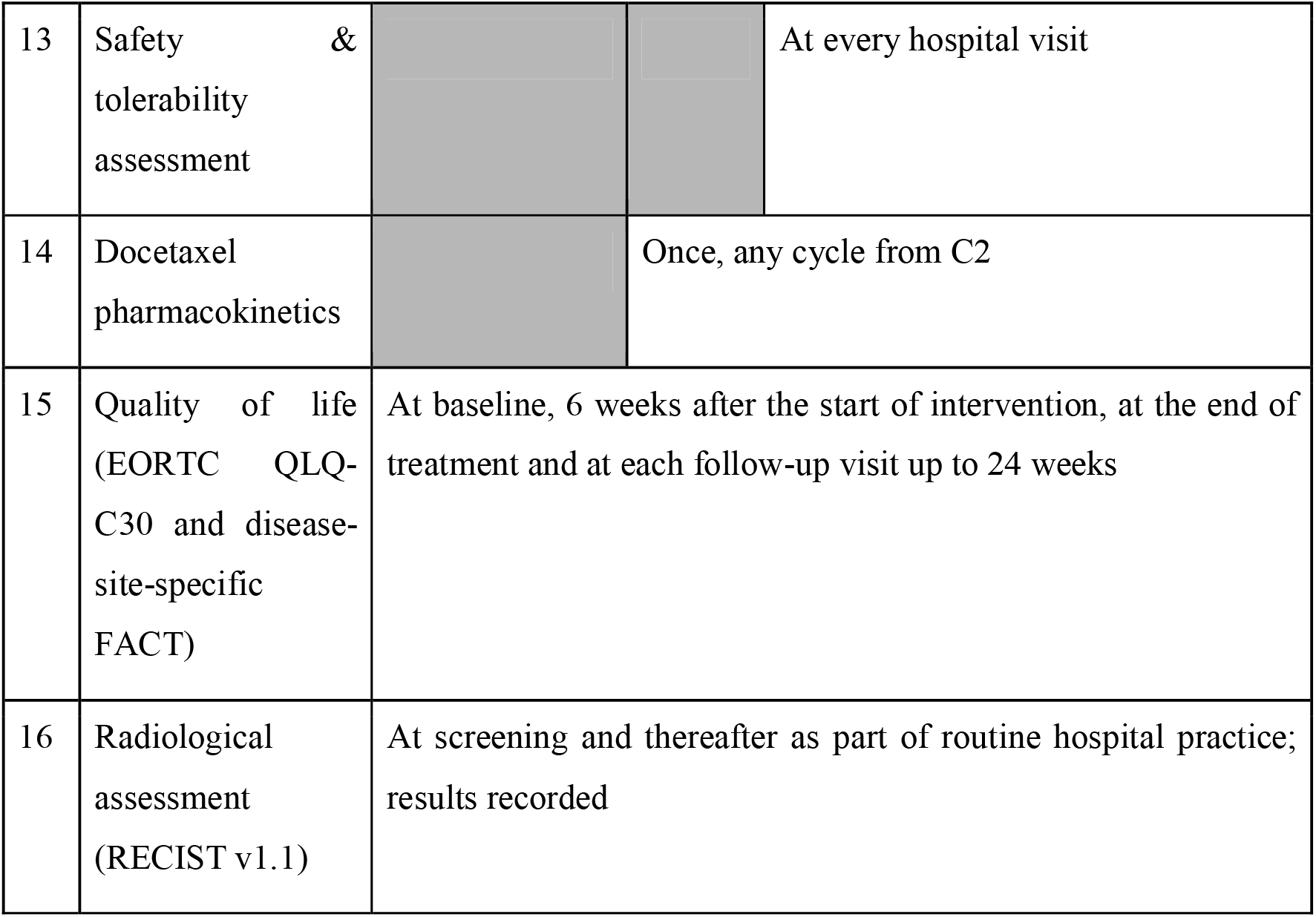
Study procedure by visit.

### Participant withdrawal

Participants were discontinued from the trial if they withdrew consent (participation was voluntary and consent could be withdrawn at any time) or for any of the following reasons: a serious adverse event; an increase in the severity of a concomitant chronic disease; a new acute illness; a non-medical event making continuation unavoidably impossible; non-compliance with the protocol; or at the discretion of the study doctors. Non-compliance was defined as taking less than 80% of study tablets, refusing the protocol investigations, not completing the questionnaires, or attending follow-up visits more than 5 days outside the schedule. A temporary hold of the study drug on the advice of the treating or Ayurvedic physician for an adverse event was not counted as discontinuation. The reason for each discontinuation was recorded. The efficacy analysis follows the intention-to-treat principle, so all randomized participants were analyzed in their allocated arm

### Consent

Eligible patients received the participant information sheet and consent form after a discussion with the investigator, and were allowed time to decide before signing. Written informed consent was signed by the participant and the investigator before enrolment. A separate consent was obtained for pharmacokinetic sampling, which was carried out only in those who agreed. Data were coded without personal identifiers and stored under the supervision of the Principal Investigator for 5 years. No participant will be identified in any publication without permission.

### Randomization, allocation and blinding

Allocation of the patients to the control arm or the treatment arm were done using stratified permuted block randomization with variable block size, in a 2:1 ratio favouring AsSET, stratified by planned cumulative docetaxel dose (≤ 225mg/m² vs. > 225mg/m²)

The original protocol specified adaptive randomisation (minimisation); this was changed to stratified permuted block randomisation by amendment (Annexure 1), because there was only one stratification factor. The change took effect on 14 November 2023 (protocol version 3.1); the five participants enrolled before that date were allocated by minimisation.

Participants and treating physicians were blinded to allocation. Study bottles were identified only by an alphanumeric code held by the study statistician, who sent the code to the central pharmacy (Clinical Pharmacology Unit pharmacy, ACTREC) to dispense the study drug after randomization. AsSET and placebo tablets were identical in colour, size, shape, texture, smell and taste, and were supplied in similar sealed HDPE bottles of 50 tablets. Each label carried the randomization code, the number of tablets, the dose, the manufacture and expiry dates, and the manufacturer’s name and address. Unblinding was permitted only for a serious adverse event requiring knowledge of the allocation, and had to be documented and reported to the sponsor. The study statistician, who held the allocation codes, and the dispensing pharmacist were not blinded. The database was locked on 17 September 2026 and the locked dataset was transferred to an independent statistician who remains blinded to treatment allocation. Unblinding has not yet been done and outcome analysis has not started.

**Figure 2:**
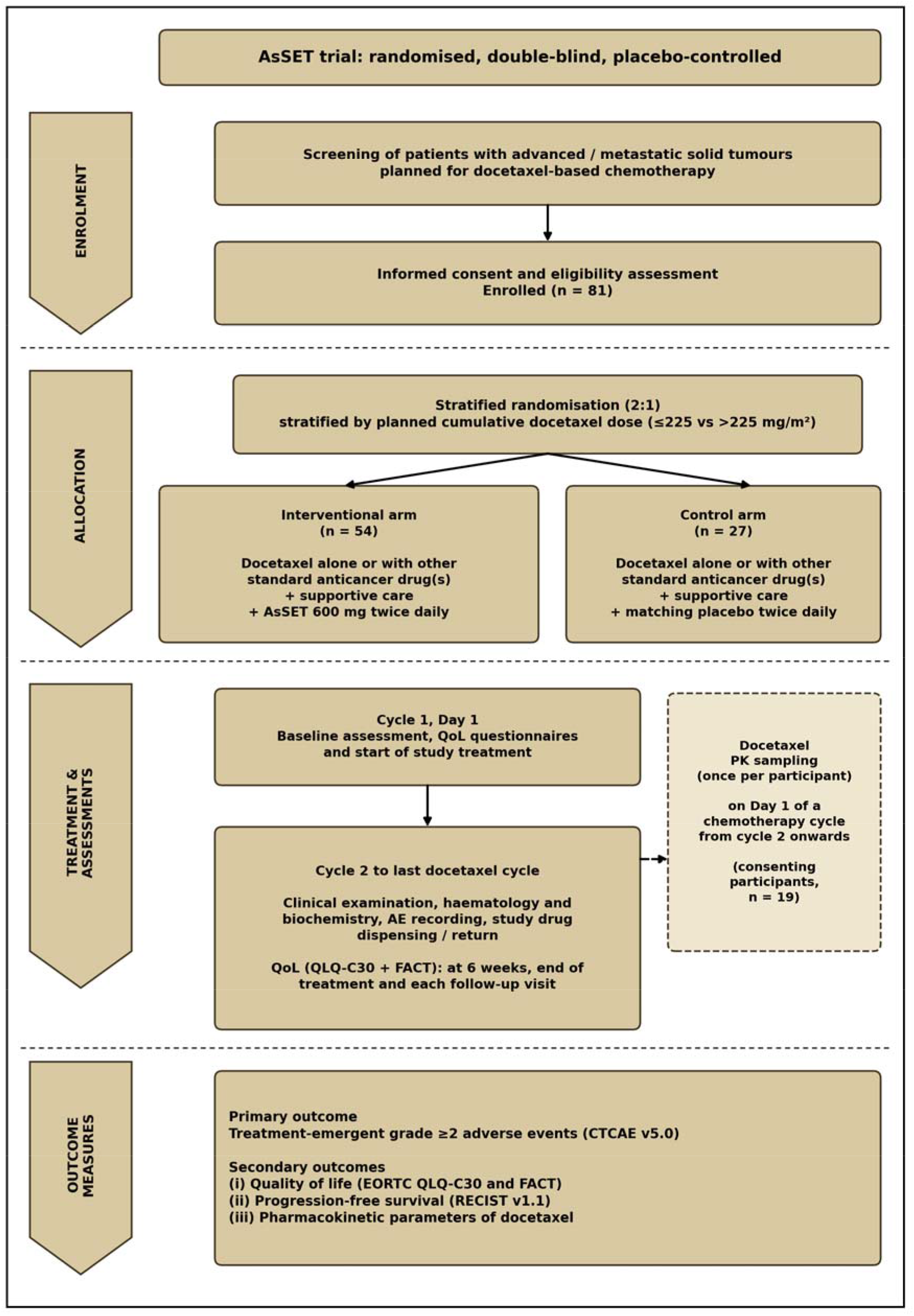
Flow chart of trial recruitment and follow-up in the AsSET Trial.

## OUTCOME MEASUREMENTS

### Primary outcome

The primary outcome is the proportion of patients experiencing at least one treatment emergent adverse events of grade ≥2 (haematological or non-haematological), graded according to CTCAE v5.0, from the first dose of study drug to study completion (up to 24 weeks). All undesirable clinical symptoms (vital signs, biochemistry, haematology) during the trial shall be reported as adverse events defined as per CTCAE v5.0.

### Secondary outcome

Quality of Life (QoL) scores and the disease-site-specific FACT scale, Progression-Free Survival (PFS), haematological and biochemical parameters; and docetaxel pharmacokinetic parameters.

Health-related quality of life (QoL) was assessed using two validated tools: the EORTC QLQ-C30 v3.0 (20), which covers functional scales (physical, role, cognitive, emotional and social), symptom scales and global health status, with scores linearly transformed to a 0–100 scale, and the disease-site-specific Functional Assessment of Cancer Therapy (FACT) scale (29), which covers physical, social/family, emotional and functional well-being. Both tools were administered at baseline, at 6 weeks from start of intervention, at the End of Treatment (EoT) and during each subsequent follow-up visit up to 24 weeks.

Progression free survival(PFS) is defined as the time from start of study treatment to the first documented progression as per RECIST v1.1 or death from any cause, whichever occurs first (Timeframe: up to 24 weeks). Pharmacokinetic analysis – All PK assays of docetaxel will be conducted in consenting participants in any chemotherapy cycle from Cycle 2 onwards. Approximately 3 mL of blood was collected in EDTA tubes at nine timepoints up to 24 hours [from the start of docetaxel infusion] (0.5 h, end of infusion, 1.17, 1.5, 2, 3, 6, 12 and 24 h). Docetaxel concentrations will be measured by a validated High Performance Liquid Chromatography(HPLC)E method (linear range 50–5000 ng/mL) (21), in the Clinical Pharmacology Laboratory, ACTREC-TMC. Docetaxel parameters (AUC0–inf, Cmax, Vd, CL, t1/2 and MRT) will be compared between arms.

### Sample size

The sample size was estimated for the primary outcome (the proportion of participants with grade ≥2 adverse events) using the Select Statistics two-proportion calculator, assuming a two-sided significance level of 0.05, 80% power and an anticipated absolute difference of 0.30 between the proportions of participants experiencing grade ≥2 adverse events in the AsSET and placebo groups. After allowing for an anticipated 10% dropout, the target sample size was 81 participants. A total of 81 participants were randomly allocated in a 2:1 ratio to docetaxel-based chemotherapy plus AsSET (n = 54) or docetaxel-based chemotherapy plus placebo (n = 27). The 2:1 allocation ratio was chosen to increase exposure to the investigational product, which had been well tolerated in earlier clinical studies, including in healthy volunteers (18).

### Study monitoring

Adverse events (AEs) were recorded and graded at every visit using CTCAE version 5.0(19), and serious adverse events (SAEs) were reported to the ethics committee and the sponsor as required by the New Drugs and Clinical Trials Rules 2019. Expected Adverse Events were mild constipation due to AsSET and, for docetaxel, neutropenia, hypomagnesaemia, acne-like rash, allergic reactions, pneumonia, mucositis, conjunctivitis, nausea and diarrhoea. Internal monitoring was done by Institutional Ethics Committee from the trial institution. External monitoring was carried out by the sponsors (Ministry of Ayush and KIHS-MRF). There was no data monitoring committee; safety was monitored through CTCAE grading at every visit and reporting of serious adverse events under the NDCT Rules 2019.

### Statistical analysis

The efficacy analysis population comprised all randomised participants (intention to treat). For the primary outcome, the proportion of participants with grade ≥2 adverse events will be compared between arms using the χ² test or Fisher’s exact test; the same tests will be used for other categorical variables. Continuous variables (including quality-of-life scores, haematological and biochemical values, and pharmacokinetic parameters) will be summarised as mean (SD) or median (interquartile range) and compared between arms using the t-test or the Mann–Whitney U test, according to the distribution of the data assessed with the Shapiro–Wilk test. Progression-free survival will be estimated by the Kaplan–Meier method and compared using a stratified log-rank test and a stratified Cox proportional hazards model (Efron’s method for ties), with treatment as the single covariate and the randomisation stratification factor as the stratifier, reporting the hazard ratio with 95% CI. All tests will be two-sided, with P < 0.05 considered significant. The statistical methods described here are those specified in the latest approved protocol (version 4.0). The database was locked on 17 September 2026, and no outcome data have been analysed by treatment arm as of the date of this manuscript.

## SAFETY AND ANCILLARY CARE

All adverse events were managed free of cost at Tata Memorial Centre, by admission or outpatient treatment as needed. All participants received reimbursement of Rs 1,500 for each non-routine visit, and for the pharmacokinetic sampling related visit. AsSET was provided until the last cycle of docetaxel.

## STUDY CONDUCT

The trial was conducted in accordance with ICH Good Clinical Practice and the New Drugs and Clinical Trials Rules 2019. Participants were identified by unique codes, with no personal identifiers disclosed. Participant data and study logs will be kept for 5 years after completion of the trial. Protocol amendments were submitted in writing to the Institutional Ethics Committee (IEC) and implemented after its approval. (list of important amendments related to trial conduct: Annexure 1)

## ETHICS AND DISSEMINATION

The results of this trial, regardless of its findings- Positive, negative or inconclusive will be reported and published with equal rigour, maintaining the transparency of research findings.

## DATA AVAILABILITY

The complete trial dataset will be made available to other research on reasonable request, following appropriate data-sharing agreements, after publications of the results.

## AUTHORS CONTRIBUTION

PS- Writing original draft, validation, analysis, editing and cover letter drafting; AS- Writing original draft, review, editing, validating, analysis; SS- data collection, review and editing; PB- data collection; AJ- review and editing; AV- study design, conceptualization; VO-review and editing; VN- review and editing; KP- review and editing; NR- review and editing; ARa- review and editing; ARt- study design, conceptualization, methodology, validation, analysis, review and editing; JJ- study design, conceptualization, methodology, validation, analysis, review and editing; VG- study design, conceptualization, methodology, validation, analysis, review and editing

## DISCUSSION

Cancer treatment typically involves multiple therapeutic modalities such as surgery, chemotherapy, immunotherapy, and hormonal or geneLbased intervention. Despite these advanced therapeutic interventions, these approaches are often limited by significant sideLeffects that compromise patient tolerance and may lead to dose reductions or treatment discontinuation (22). Docetaxel is a taxaneLbased chemotherapeutic and cytotoxic drug which is approved for breast, non-small cell lung, prostate, gastric, and head and neck cancers (7). Its mechanisms involve stabilisation of microtubules, arresting cells in the G2/M phase, and thereby inhibiting tumour growth (23,24). DoseLlimiting toxicities of docetaxel include shortLlasting neutropenia, alopecia, mucositis, neuropathy, fluid retention, and hypersensitivity reactions that markedly affect quality of life(23–25).

Given these challenges, there is a pressing need for, and growing interest in developing adjunctive therapies that would minimize the negative impacts of drugs used in cancer chemotherapy treatment while preserving its efficacy(22,26). Ayurvedic medicines offer several potential agents with proven biological activity. We are investigating an addLon strategy using a aqueous standardized extract of Tinospora cordifolia (AsSET), which has reported immunomodulatory and chemoprotective properties(8–14) and a favorable safety profile (8,18). This randomized, doubleLblind, placeboLcontrolled trial aims to assess clinical benefits of Guduchi and evaluate whether combining docetaxel with AsSET can reduce chemotherapyLrelated adverse events, thus improving tolerability, and enhancing quality of life in patients with advanced, recurrent or metastatic solid tumours.

The design addresses limitations of earlier clinical evaluations of T. cordifolia as a chemoprotective agent. These were small, one lacked between-group comparisons and CONSORT-compliant reporting, and the placebo-controlled study in breast cancer was reported only as a postgraduate thesis in a different chemotherapy setting (15,16). The use of a aqueous standardized extract manufactured to defined batch specifications supports batch consistency and reproducibility, which is important for herbal intervention trials. The pharmacokinetic sub-study addresses the clinically important question of a possible herb–drug interaction between AsSET and docetaxel, which has so far been examined only in preliminary in vitro studies (27,28).

## CONCLUSION

To our knowledge, this trial is the first prospectively registered, randomized controlled trial to evaluate the chemoprotective efficacy of an aqueous standardized extract of T. cordifolia as an adjunct to docetaxel-based chemotherapy. If the findings demonstrate a significant reduction in chemotherapy-induced adverse events without attenuation of docetaxel pharmacokinetics or antitumor activity of docetaxel, they would generate robust evidence to support the integration of AsSET into oncology practice. Results will be reported transparently, whether positive, negative, or inconclusive, to contribute to the evidence on herbal adjuncts in cancer care.

## TRIAL STATUS

The manuscript is based on Protocol Version 4.0, (24 February 2026). The protocol has been approved by the Institutional Ethics Committee (IEC) of Advanced Centre for Treatment, Research and Education in Cancer (ACTREC), Tata Memorial Centre (TMC), Navi Mumbai. Version 3.0 (initial approval on 28 June 2023); version 3.1, dated 14 November 2023, introduced the amendments to the eligibility criteria, the randomisation method and the sponsor details listed in Annexure 1; version 4.0, dated 24 February 2026, revised the budget only and did not change the conduct of the trial. The trial is registered with CTRI-CTRI/2023/08/056129. Participant recruitment began on 2 November 2023. Currently, we have successfully completed the enrollment of all the 81 patients. Nineteen participants consented to pharmacokinetic sampling. The database was locked on 17 September 2026 and the locked dataset was transferred to an independent statistician who remains blinded to treatment allocation. Unblinding has not yet been done and outcome analysis has not started.

## DECLARATION OF GENERATIVE AI IN SCIENTIFIC WRITING

We used Claude Sonnet 5.5 (26 October 2026, Anthropic, San Francisco, CA, USA) to correct grammar and punctuations as well for paraphrasing the original text of protocol. The authors reviewed and edited the content as needed, and take full responsibility for the content of the publication.

## Supporting information

Annexure 1

## Data Availability

All data produced in the present work are contained in the manuscript

http://ctri.nic.in

## ACKNOWLEDGEMENT

The authors thank Dr. Pankaj Chaturvedi, Director, ACTREC-TMC for permitting and supporting the conduct of the study. We thank the consulting physicians and resident doctors of ACTREC-TMC and Tata Memorial Hospital for their support and for referring patients to the study. We are grateful to the Ministry of Ayush, Government of India, and the Kasturba Integrative Health Sciences – Medical Research Foundation (KIHS-MRF) for funding and guidance throughout, and to Pharmanza Herbal Pvt. Ltd. for supplying the investigational products (AsSET and matching placebo). Above all, we thank the patients who consented to take part and cooperated with us throughout the study.

## Abbreviation

AsSET: Aqueous Standardized Extract of Tinospora cordifolia
CTCAE: Common Terminology Criteria for Adverse Events
PFS: Progression-free survival
GM-CSF: Granulocyte macrophage-colony stimulating factor
GFR: Glomerular Filtration rate
AUC□-inf: Area Under the (concentration-time) Curve from time zero to infinity
AE: Adverse event
ECOG: Eastern Cooperative Oncology Group
EORTC QLQ-C30: European Organisation for Research and Treatment of Cancer Quality of Life Questionnaire – Core 30
FACT: Functional Assessment of Cancer Therapy
IEC: Institutional Ethics Committee
KIHS-MRF: Kasturba Integrative Health Sciences – Medical Research Foundation
PK: Pharmacokinetic
QoL: Quality of life
RECIST: Response Evaluation Criteria in Solid Tumors
SAE: Serious adverse event

