## Supplementary material for "Study protocol for a randomized, double-blind, placebo-controlled clinical trial to assess chemoprotective efficacy of Aqueous standardized extract of *Tinospora cordifolia* (AsSET) in patients receiving Docetaxel based chemotherapy for advanced / metastatic solid tumors": Annexure 1

**Amendments to the trial protocol that relate to the conduct of the trial**

**Protocol version history**

Initial protocol: approved by the Institutional Ethics Committee (IEC), ACTREC, Tata Memorial Centre, on 28 June 2023 (version 3.0). The first participant was enrolled on 2 November 2023, under this protocol.

Version 3.1, dated 14 November 2023: introduced the four amendments listed in the table below (secondary sponsor, inclusion criteria, exclusion criteria and method of generating the random sequence). The five participants enrolled before this date were enrolled and allocated under the initial protocol (minimisation).

Version 4.0, dated 24 February 2026: revised only budget section. It made no change to eligibility, interventions, assessments, outcomes, randomisation or analysis, and is therefore not listed in the table.

**Amendments introduced in version 3.1**

| **Sr. No.** | **Section** | **Old text** | **Update/ Modification** | **Justification** |
| --- | --- | --- | --- | --- |
| 1. | Secondary Sponsor | Nil | Name: Kasturba Integrative Health Sciences- Medical Research Foundation (KIHS-MRF)  Address: 5th & 6th floor, Jain Aaradhana Dham Bldg.,17th KD Road, Vile Parle (west), Mumbai 400056 | At the time of CTRI registration, the proposal was submitted to ICMR for funding. As the proposal was not accepted by ICMR, it was subsequently submitted to the Ministry of AYUSH, Government of India. The study initially commenced  following IEC approval.  As we did not receive any funds from ICMR, our collaborator (Secondary sponsor) Kasturba Integrative Health Sciences- Medical Research Foundation (KIHS-MRF) provided us with immediate partial funding for smooth conduct of the trial in the initial phase of the study. Later when we received the grants from our primary sponsor, the other expenses towards the study were met. |
| 2. | Inclusion criteria | 1. Voluntarily signed informed consent. 2. Patients with evaluable disease, histologically documented confirmed diagnosis of solid malignancy of Head & Neck cancer, Lung cancer, and Prostate cancer. 3. Chemo-naive for locally advanced / metastatic disease; recurrent / metastatic squamous cell carcinoma of head & neck (R/M SCCHN), Non-small cell Lung cancer (NSCLC) and metastatic castration resistant prostate cancer (mCRPC). 4. Males and females, 18 to 65 years of age (both inclusive). 5. Eastern Cooperative Oncology Group (ECOG) performance status ≤ 2. 6. Adequate bone marrow reserve (WBC at least 3,000/mm^3^, neutrophil count ≥2000/mm^3^, platelet count ≥1,00,000mm^3^ and haemoglobin level≥ 8.0 g/dL), renal function (GFR≥50mL/min), liver function [total bilirubin level ≤2 times upper normal limit (UNL) and serum transaminases levels ≤2.5 times UNL/ ≤5 times for liver metastasis and/or obstructive jaundice. 7. Life expectancy minimum 12 weeks. 8. Patients willing to practice birth control during and for 2 months after treatment. 9. Subjects who have tested negative for HIV, Haigh and pregnancy. 10. Subjects who have received the vaccination for COVID-19 at least first two doses. | 1. Voluntarily signed informed consent. 2. Patients with evaluable disease, histologically documented confirm diagnosis of solid malignancy. 3. Males and females, 18 to 65 years of age (both inclusive). 4. Eastern Cooperative Oncology Group (ECOG) performance status ≤ 2. 5. Adequate bone marrow reserve (WBC at least 3,000/mm^3^, neutrophil count ≥2000/mm^3^, platelet count ≥1,00,000mm^3^ and haemoglobin level≥ 8.0 g/dL), renal function (GFR≥50mL/min), liver function [total bilirubin level ≤2 times upper normal limit (UNL) and serum transaminases levels ≤2.5 times UNL/ ≤5 times for liver metastasis and/or obstructive jaundice. 6. Life expectancy minimum 12 weeks. 7. Patients willing to practice birth control during and for 2 months after treatment. 8. Subjects who have tested negative for HIV, HBsAg and HCV. | **Criteria no. 2 has been revised, no. 3 has been removed**  **Reason:** The primary objective is to study efficacy of AsSET drug in patients receiving docetaxel-based chemotherapy so we wish to enrol patients who have previously received any chemotherapy except docetaxel to fasten the accrual of the study. Additionally, including this patient population in the study will not affect our primary objective  **Criteria no. 10 has been removed**  **Reason**: This protocol was written in 2020 when there was wave of COVID-19. So as precautionary measure it was included in eligibility criteria. But in year 2023 while making amendments to IEC, majority of the population was vaccinated and the situation was in a better phase so we wish to not keep this criteria as mandatory for eligibility. |
| 3. | Exclusion criteria | 1. Patients who have received prior treatment with docetaxel. 2. Patients with major illness including active cardiac, hepatic, endocrine, renal or psychiatric disorders not adequately controlled with current therapy. 3. Patients with brain metastases or primary CNS malignancies. 4. Any condition, including the presence of laboratory abnormalities, which places the subject at unacceptable risk if he/she were to participate in the study, and which may obscure the evaluation of toxicity or alters drug metabolism. 5. Patients who have received anti-tumor treatments such as radiotherapy, targeted therapy, immunotherapy, and other clinical study drugs within 4 weeks before the first dose of the investigational drug. 6. Traditional medicines/natural product with anti-tumor indications and/or cancer chemotherapy toxicity resolving potential within 4 weeks before the first dose of the investigational drug. 7. Patients who have received corticosteroid (prednisone > 10 mg/day or equivalent) or other immunosuppressive therapies within 4 weeks before the first dose of the investigational drug. 8. Covid vaccination in the preceding 4 weeks or COVID positive by RT-PCR and who have received any live attenuated vaccine within 4 weeks before the first dose of the investigational drug or planned to receive live attenuated vaccine during the study period. 9. Patients who have undergone major surgery within 4 weeks before the first dose of the investigational drug, or who need to undergo elective surgery during the immediate subsequent period of the study. 10. Patients who are pregnant or lactating. | 1. Patients who have received prior treatment with docetaxel. 2. Patients with major illness including active cardiac, hepatic, endocrine, renal or psychiatric disorders not adequately controlled with current therapy. 3. Patients with brain metastases or primary CNS malignancies. 4. Any condition, including the presence of laboratory abnormalities, which places the subject at unacceptable risk if he/she were to participate in the study, and which may obscure the evaluation of toxicity or alters drug metabolism. 5. Patients who have received anti-tumor treatments such as radiotherapy, targeted therapy, immunotherapy, and other clinical study drugs within 4 weeks before the first dose of the investigational drug. 6. Traditional medicines/natural product with anti-tumor indications and/or cancer chemotherapy toxicity resolving potential within 4 weeks before the first dose of the investigational drug. 7. Patients who have received corticosteroid (prednisone > 10 mg/day or equivalent) or other immunosuppressive therapies within 4 weeks before the first dose of the investigational drug. 8. Patients who have received any live attenuated vaccine within 4 weeks before the first dose of the investigational drug or planned to receive live attenuated vaccine during the study period. 9. Patients who have undergone major surgery within 4 weeks before the first dose of the investigational drug, or who need to undergo elective surgery during the immediate subsequent period of the study. 10. Patients who are pregnant or lactating. | **Criteria no. 8 has been revised**  **Reason**: We have removed the criteria of mandatory COVID vaccination. This protocol was written in 2020 when there was wave of COVID-19. So as precautionary measure it was included in eligibility criteria. But in year 2023 while making amendments to IEC, majority of the population was vaccinated and the situation was in a better phase so we wish to not keep this criteria as mandatory for eligibility. |
| 4. | Method of Generating Random Sequence | Adaptive randomization, such as minimization | Stratified permuted block randomization technique with variable block size 2 or 4 | As the trial has only one stratification factor, stratified permuted block randomisation is more efficient than minimisation. |
